# The impact of Type 2 diabetes in Parkinson’s disease

**DOI:** 10.1101/2021.10.21.21265308

**Authors:** Dilan Athauda, James Evans, Anna Wernick, Gurvir Virdi, Minee Liane-Choi, Michael Lawton, Nirosen Vijiaratnam, Christine Girges, Yoav Ben-Shlomo, Khalida Ismail, Huw Morris, Donald Grosset, Thomas Foltynie, Sonia Gandhi

## Abstract

**Importance:** Type 2 diabetes (T2DM) is an established risk factor for developing Parkinson’s disease (PD) but its effect on disease progression is not well understood.

**Objective:** To examine the effects of co-morbid T2DM on Parkinson’s disease progression and quality of life.

**Design:** We analysed data from the *Tracking Parkinson’s* study, a large multi-centre prospective study in the UK.

**Participants:** The study included 1930 adults with recent onset PD, recruited between February 2012 and May 2014, and followed up regularly thereafter.

**Exposure:** A diagnosis of pre-existing T2DM was based on self-report at baseline. After controlling for confounders, an evaluation of how T2DM affects PD was performed by comparing symptom severity scores; and analyses using multivariable mixed models was used to determine the effects of T2DM on Parkinson’s disease progression.

**Main Outcomes and Measures:** The impact of T2DM on Parkinson’s disease severity was derived from scores collected using the Movement Disorders Society Unified Parkinson’s Disease Rating Scale (MDS-UPDRS), Non-Motor Symptoms Scale (NMSS), Montreal Cognitive Assessment (MoCA), Questionnaire for impulsive-compulsive disorders in PD (QUIP), Leeds Anxiety and Depression Scale (LADS), and Schwab and England ADL scale.

**Results:** We identified 167 (8.7%) patients with PD and T2DM (PD+T2DM) and 1763 (91.3%) with PD without T2DM (PD). Patients with T2DM had more severe motor symptoms, as assessed by MDS-UPDS III 25.8 (0.9) vs 22.5 (0.3) p=0.002, had significantly faster motor symptom progression over time (p=0.012), and T2DM was an independent predictor for the development of substantial gait impairment (HR 1.55, CI 1.07-2.23, p=0.020). Patients were more likely to have loss of independence (OR 2.08, CI 1.34-3.25, p=0.001); and depression (OR 1.62, CI 1.10-2.39, p=0.015), and developed worsening mood (p=0.041) over time compared to the PD group. T2DM was also an independent predictor for the development mild cognitive impairment (HR 1.7, CI 1.24-2.51, p=0.002) over time

**Conclusions and relevance:** T2DM is associated with faster disease progression in PD, highlighting an interaction between these two diseases. As it is a potentially modifiable, metabolic state, with multiple peripheral and central targets for intervention, it may represent a target for ameliorating parkinsonian symptoms, and progression to disability and dementia.

**Key points:** *Question:* What is the impact of Type 2 diabetes on Parkinson’s disease progression?

*Findings:* In this prospective study of 1930 patients with recent onset PD, T2DM is an independent risk factor for more severe motor features, non-motor symptoms, and poorer quality of life; and importantly is associated with faster motor and non-motor symptom progression, and increases the risk of developing cognitive impairment.

*Meaning:* T2DM is identified as a new factor that alters Parkinson’s disease progression. T2DM predicts both motor and non-motor symptom progression in PD and is associated with poorer quality of life, highlighting an interaction of two chronic disease states. This highlights a particular need for improved treatment in this subgroup of patients with Parkinson’s.

## Introduction

Parkinson’s disease (PD) affects 6 million people worldwide and its prevalence is expected to increase in response to an ageing population^1^. While ageing is undoubtedly the most important risk factor, there is evidence that Type 2 diabetes (T2DM) is a modest risk factor^2^, with studies suggesting diabetic patients with a long disease duration and more severe complications are at the greatest risk of developing PD^3^. Additionally, accumulating evidence suggests these diseases share common biological mechanisms (reviewed here^4^) and shared genetic links^5^, which highlights dysfunctional insulin signalling as a possible convergent pathway between these conditions^6^.

Beyond its effects on PD risk, some studies have suggested co-morbid T2DM in patients with PD may influence disease progression and observed: accelerated motor progression^7,8^, reduced time to develop levodopa-related motor complications^9^, gait difficulties ^10,11^ and cognitive impairment ^12,13^, in comparison to PD patients without T2DM. However, generalizability from these studies has been limited by the relatively small numbers of patients included with co-morbid T2DM.

Utilising the *Tracking Parkinson’s* cohort, a long-term observational study into PD, our aims were to 1) evaluate the association of co-morbid T2DM on PD severity in patients recently diagnosed with PD, and 2) determine whether T2DM negatively affects disease progression. In view of accumulating data suggesting anti-glycaemic treatments may be useful in the treatment of PD, we also conducted an exploratory analysis to determine 3) if metformin use confers any protective effects on the severity and long-term outcomes.

## Methods

### Study design and data collection

Data from *Tracking Parkinson’s* including demographic, clinical, imaging, and biospecimen measures, that have been collected for more than six years were analysed. The study set-up and design have been previously reported^14^. Enrolled participants were recruited with a clinical diagnosis of PD fulfilling UK Brain Bank criteria and included both drug-naïve and treated patients, aged 18–90 years. Recent onset cases were diagnosed with PD in the preceding 3.5 years, and recruitment was completed between February 2012 and May 2014.

The following features were collected: demographics, diagnostic features at presentation, ethnicity, education, medication history, body mass index (BMI) and comorbidities. A concurrent diagnosis of pre-existing T2DM was based on self-report at baseline. L-dopa equivalent daily dose (LEDD) was calculated using an established formula^15^.

### Outcomes

We used several motor and non-motor features previously used in other studies as “severe/advanced” disease markers or clinical milestones^16^, and used them to identify their appearance at baseline entry into the study (the reasoning that, even at a short disease duration, patients with a greater number of these markers at baseline could be identified as having a more severe disease phenotype); and also used them to monitor long term disease progression (See Supplementary data for further details on outcome calculations).

### Statistical analyses

Group comparisons between PD and PD+T2DM groups were performed at baseline, using multivariate analysis of covariance with post hoc Bonferroni correction. Categorical variables were compared using Fisher’s exact test and multivariate logistic regression was used to determine the adjusted odds ratios (OR). Potential confounders were included the statistical models guided by mechanisms proposed and depicted in directed acyclic graphs (DAGs) which considers each variable in relation to the exposure and outcome, as both the failure to adjust for a confounder, and over-adjusting for an intermediate variable can lead to biased results^17^ (Supplementary Material; Figure 1). The minimally sufficient adjustment set for estimating the total effect of T2DM on PD severity highlighted age, sex, ethnicity and BMI as covariates. Additionally, we included PD duration, vascular risk factors, Hoehn & Yahr stage and LEDD as covariates.

**Figure 1:**
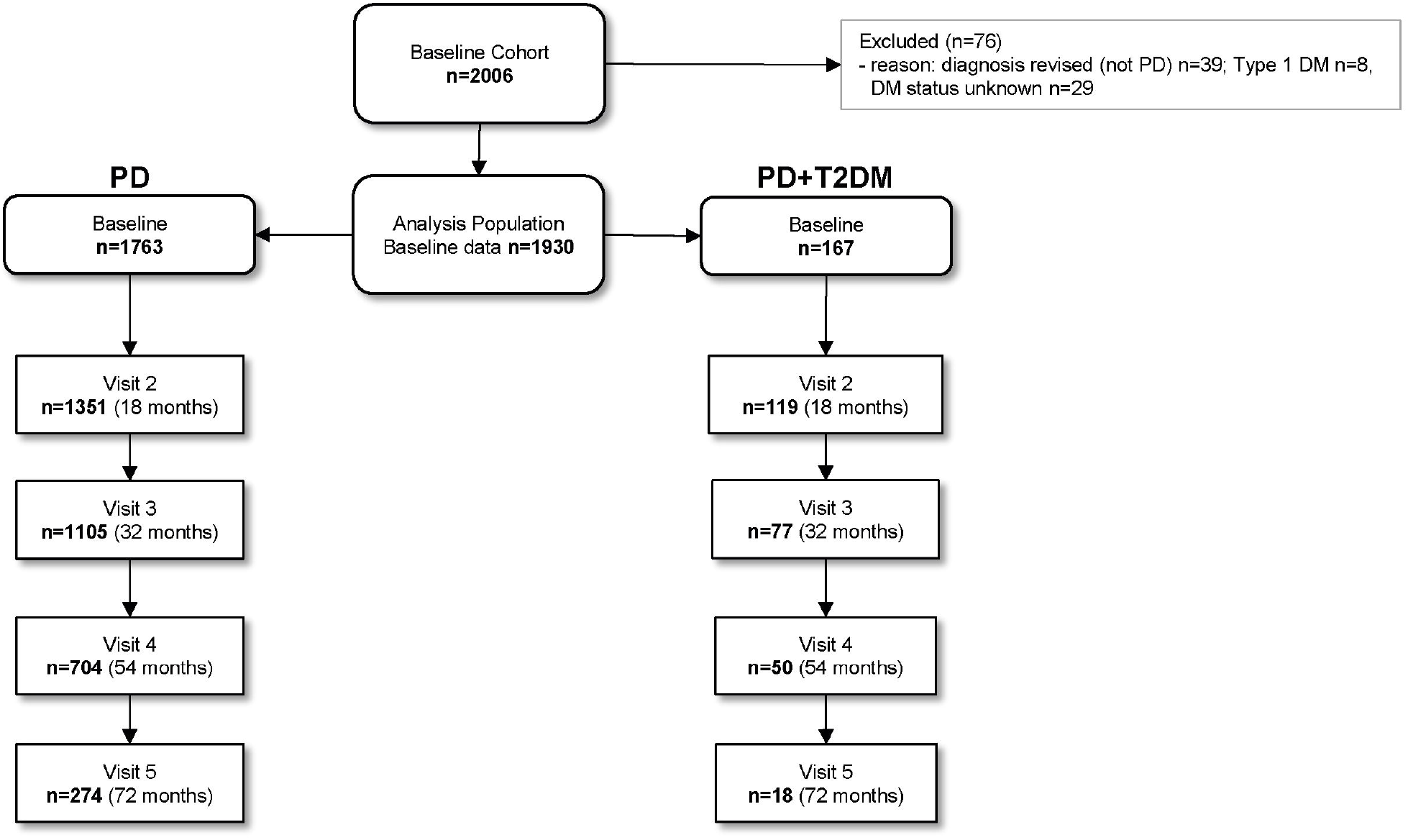
Flow diagram of the study population

For longitudinal analysis, we performed a survival analysis to determine if T2DM influenced PD progression based on the appearance of clinical milestones, as previously defined. Kaplan-Meier survival curves were plotted and log-rank tests performed to evaluate whether the presence of T2DM could predict disease progression. The analyses were repeated including age, sex, ethnicity, PD duration, baseline Hoehn & Yahr stage, LEDD, vascular risk factors and BMI as covariates. Only the time to occurrence of the first event in a category for a given subject was used in the subsequent Cox regression model. Participants at baseline who had already developed the clinical milestone (outcome) were excluded from the model.

To assess the effect of T2DM on rate of change of a given symptom (e.g. MDS-UPDRS III score), separate linear mixed effects models with robust variance estimates were used, with examination of the interaction effects of group (PD vs T2DM) and time. Symptom progression was modelled adjusting for age, sex, disease duration, ethnicity, baseline LEDD and the baseline variable value as fixed effects for effects on the intercept and slope. Participant-specific random effects were included as both a random intercept and a random slope to account for the correlation in repeated measurements within the same participant. Due the increasing number of patient drop-outs over the follow up period, we chose a cut-off of 50% missing data as the upper threshold and subsequently chose Visit 3 as the “end-point” of the study, representing a mean follow up time of 37.8 (SD 4.3) months since study entry. Analyses were performed using SPSS statistical software, version 26.0 (IBM Corp).

## Results

### Characteristics of cohort at baseline

There were 2006 individuals recruited in the *Tracking Parkinson’s* cohort. Of those, 76 (3.8%) were excluded due to: a change in diagnosis after recruitment (n=39); if they had T1DM (n=8) or the DM status was missing (n=29) (Figure 1).

The main analysis group consisted of 1930 individuals, of whom 167 (8.7%) had co-morbid T2DM (PD+T2DM) and 1,763 (91.3%) did not (PD). Demographic features and clinical features are summarised in Table 1.

**Table 1:**
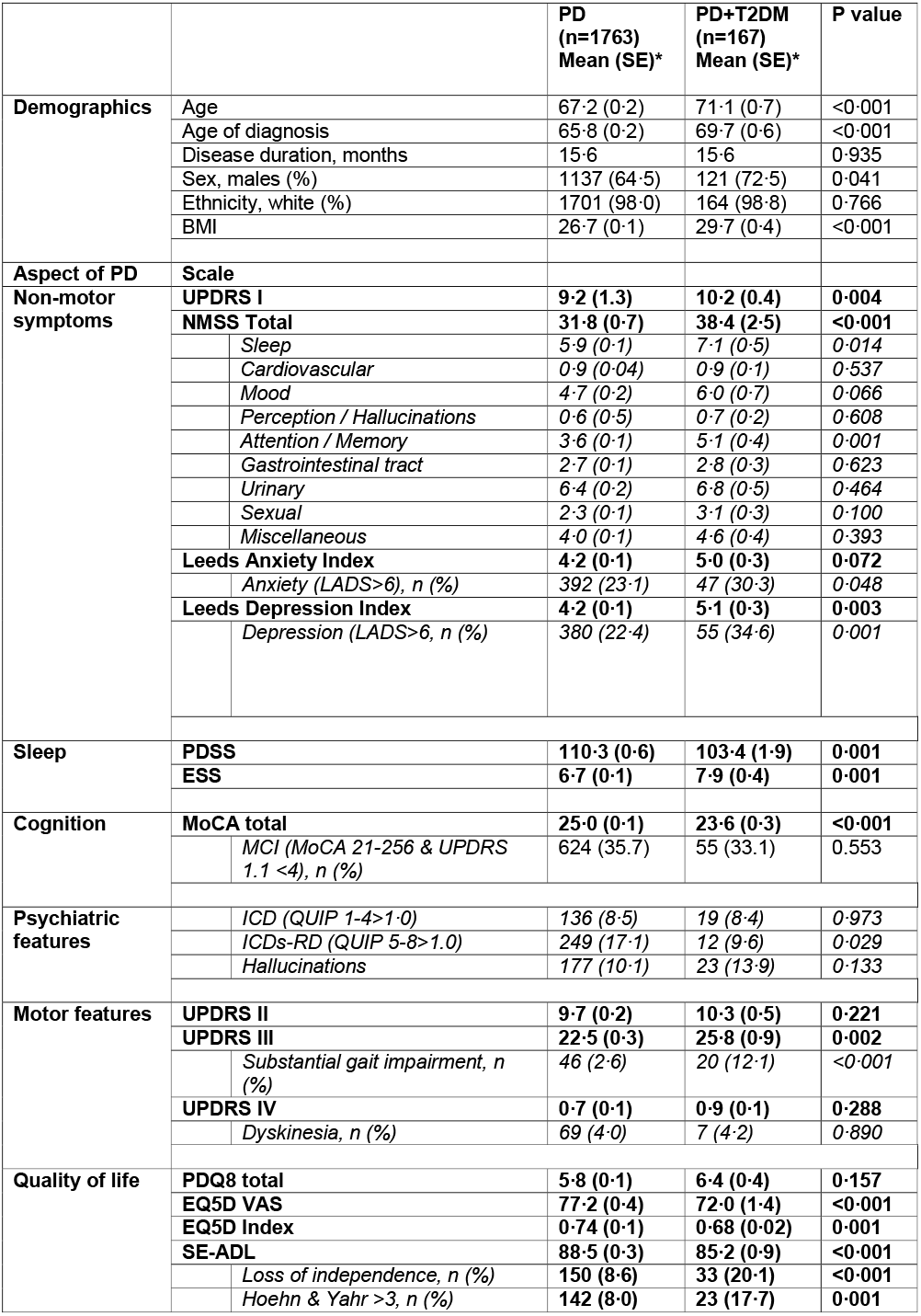
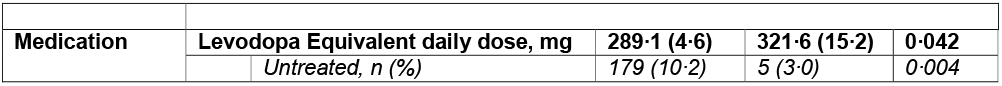
Early clinical features in PD cases without T2DM, compared to cases with T2DM.

|  |  | **PD**<br>**(n=1763)**<br>**Mean (SE)\*** | **PD+T2DM**<br>**(n=167)**<br>**Mean (SE)\*** | **P value** |
| --- | --- | --- | --- | --- |
| **Demographics** | Age | 67.2 (0.2) | 71.1 (0.7) | <0.001 |
|  | Age of diagnosis | 65.8 (0.2) | 69.7 (0.6) | <0.001 |
|  | Disease duration, months | 15.6 | 15.6 | 0.935 |
|  | Sex, males (%) | 1137 (64.5) | 121 (72.5) | 0.041 |
|  | Ethnicity, white (%) | 1701 (98.0) | 164 (98.8) | 0.766 |
|  | BMI | 26.7 (0.1) | 29.7 (0.4) | <0.001 |
| **Aspect of PD** | **Scale** |  |  |  |
| **Non-motor**<br>**symptoms** | **UPDRS I** | **9.2 (1.3)** | **10.2 (0.4)** | **0.004** |
|  | **NMSS Total** | **31.8 (0.7)** | **38.4 (2.5)** | **<0.001** |
|  | Sleep | 5.9 (0.1) | 7.1 (0.5) | 0.014 |
|  | Cardiovascular | 0.9 (0.04) | 0.9 (0.1) | 0.537 |
|  | Mood | 4.7 (0.2) | 6.0 (0.7) | 0.066 |
|  | Perception / Hallucinations | 0.6 (0.5) | 0.7 (0.2) | 0.608 |
|  | Attention / Memory | 3.6 (0.1) | 5.1 (0.4) | 0.001 |
|  | Gastrointestinal tract | 2.7 (0.1) | 2.8 (0.3) | 0.623 |
|  | Urinary | 6.4 (0.2) | 6.8 (0.5) | 0.464 |
|  | Sexual | 2.3 (0.1) | 3.1 (0.3) | 0.100 |
|  | Miscellaneous | 4.0 (0.1) | 4.6 (0.4) | 0.393 |
|  | **Leeds Anxiety Index** | **4.2 (0.1)** | **5.0 (0.3)** | **0.072** |
|  | Anxiety (LADS>6), n (%) | 392 (23.1) | 47 (30.3) | 0.048 |
|  | **Leeds Depression Index** | **4.2 (0.1)** | **5.1 (0.3)** | **0.003** |
|  | Depression (LADS>6, n (%)) | 380 (22.4) | 55 (34.6) | 0.001 |
| **Sleep** | **PDSS** | **110.3 (0.6)** | **103.4 (1.9)** | **0.001** |
|  | **ESS** | **6.7 (0.1)** | **7.9 (0.4)** | **0.001** |
| **Cognition** | **MoCA total** | **25.0 (0.1)** | **23.6 (0.3)** | **<0.001** |
|  | MCI (MoCA 21-25 & UPDRS 1.1 <4), n (%) | 624 (35.7) | 55 (33.1) | 0.553 |
| **Psychiatric**<br>**features** | ICD (QUIP 1-4>1.0) | 136 (8.5) | 19 (8.4) | 0.973 |
|  | ICDs-RD (QUIP 5-8>1.0) | 249 (17.1) | 12 (9.6) | 0.029 |
|  | Hallucinations | 177 (10.1) | 23 (13.9) | 0.133 |
| **Motor features** | **UPDRS II** | **9.7 (0.2)** | **10.3 (0.5)** | **0.221** |
|  | **UPDRS III** | **22.5 (0.3)** | **25.8 (0.9)** | **0.002** |
|  | Substantial gait impairment, n (%) | 46 (2.6) | 20 (12.1) | <0.001 |
|  | **UPDRS IV** | **0.7 (0.1)** | **0.9 (0.1)** | **0.288** |
|  | Dyskinesia, n (%) | 69 (4.0) | 7 (4.2) | 0.890 |
| **Quality of life** | **PDQ8 total** | **5.8 (0.1)** | **6.4 (0.4)** | **0.157** |
|  | **EQ5D VAS** | **77.2 (0.4)** | **72.0 (1.4)** | **<0.001** |
|  | **EQ5D Index** | **0.74 (0.1)** | **0.68 (0.02)** | **0.001** |
|  | **SE-ADL** | **88.5 (0.3)** | **85.2 (0.9)** | **<0.001** |
|  | Loss of independence, n (%) | 150 (8.6) | 33 (20.1) | <0.001 |
|  | Hoehn & Yahr >3, n (%) | 142 (8.0) | 23 (17.7) | 0.001 |

|  |  |  |  |  |  |
| --- | --- | --- | --- | --- | --- |
| Medication | Levodopa Equivalent daily dose, mg |  | 289.1 (4.6) | 321.6 (15.2) | 0.042 |
|  |  | Untreated, n (%) | 179 (10.2) | 5 (3.0) | 0.004 |

### Impact of T2DM in patients recently diagnosed with PD

Despite a similar disease duration of 15.6 months, patients with T2DM had consistently more severe symptomatology in most aspects of PD (Table 1), including: significantly worse overall non-motor symptoms, as assessed by total MDS-UPDRS I; 10.2 (SD 0.4) vs 9.2 (SD 0.1) p=0.004; NMSS 38.4 (2.5) vs 31.8 (0.7) p<0.001; worse sleep scores as assessed by Parkinson’s Sleep Scale (PDSS) 103.4 (1.9) vs 110.3 (0.6) p=0.001; Epworth Sleepiness scale 7.9 (0.4) vs 6.7 (0.1) p=0.001; worse cognitive scores assessed by MoCA 23.6 (0.3) vs 25.0 (0.1) <0.001; more severe motor symptoms, as assessed by MDS-UPDS III 25.8 (0.9) vs 22.5 (0.3) p=0.002; and worse overall quality of life scores as assessed by the EQ5D VAS 72.0 (1.4) vs 77.2 (0.4) p<0.001; EQ5DIndex 72.0 (1.4) vs 77.2 (0.2) p=0.001; and reported more impaired scores on the Schwab and England ADL scale 85.2 (0.9) vs 88.5 (0.3) p<0.001; compared to people without T2DM. Also, despite adjusting for differences in disease severity, patients with T2DM had a higher LEDD usage than participants without T2DM 321.6mg (15.2) vs 289.1mg (4.6) p=0.042.

There were significantly greater numbers of patients with T2DM who had substantial gait impairment, 12.6% vs 2.6% (p<0.001); depression, 34.6% vs 22.4% (p=0.001); and self-reported loss of independence, 20.1% vs 8.6% (p<0.001) in comparison to patients without T2DM (Table 1). Patients with T2DM also had less impulse control disorder-related behaviours (ICDs-RD) than patients without T2DM, 9.6% vs 17.1% (p=0.029).

A multivariate binomial regression analysis revealed that the T2DM was significantly and independently associated with greater gait impairment (OR 2.91, 95% CI 1.46-5.79, p=0.002), depression (OR 1.62, CI 1.10-2.39, p=0.015), and loss of independence (OR 2.08, CI 1.34-3.25, p=0.001) relative to the PD group, after adjusting for age, sex, disease duration, ethnicity, vascular risk factors, LEDD, H&Y stage and BMI (Figure 2 and Table 2).

**Figure 2:**
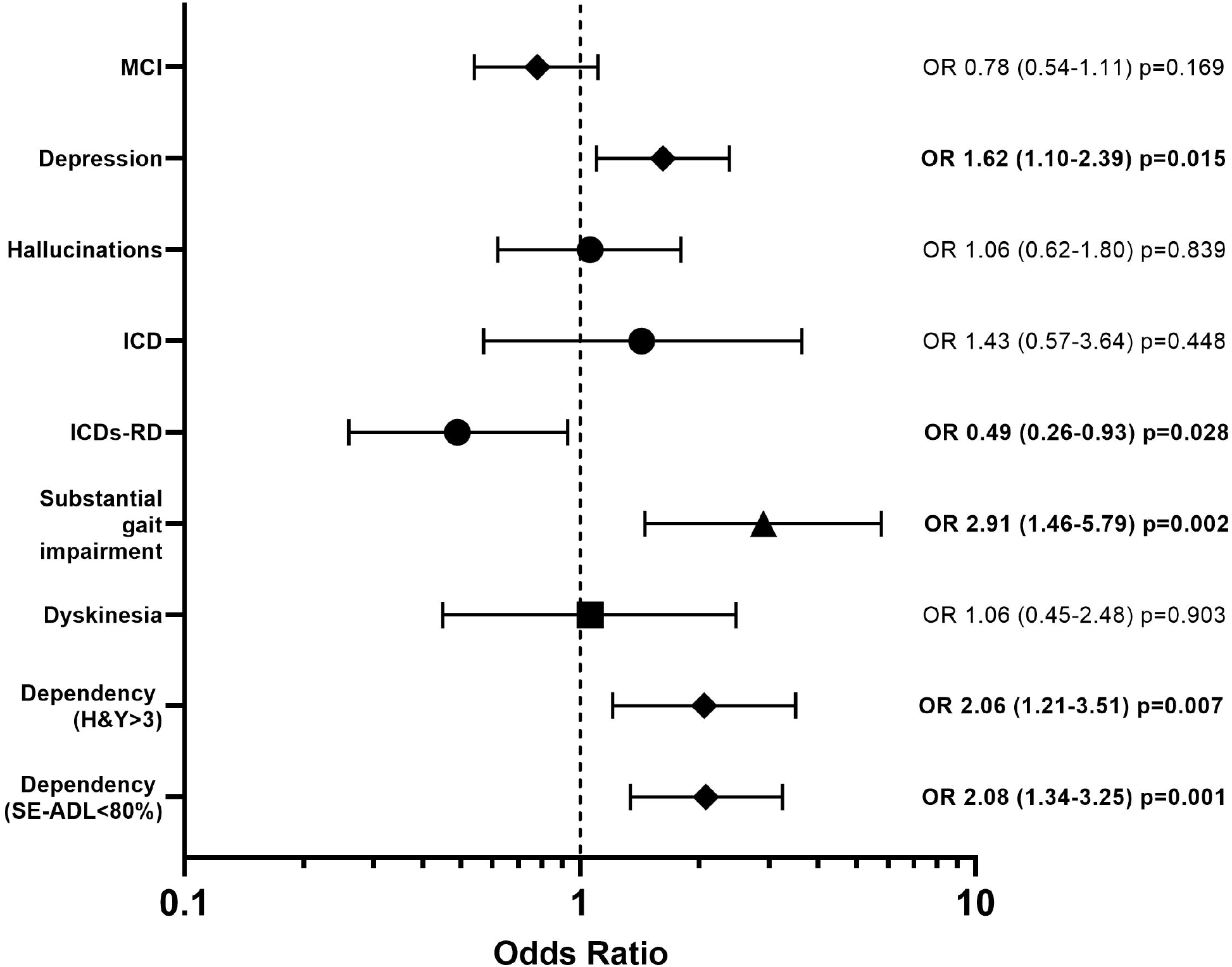
Likelihood of complications or reaching disease milestones in PD patients, according to T2DM status. Patients with T2DM were signficantly more likely to have depression, substantial gait impairment and loss of independence, and signficantly less likely to have dopamine dysregulation, than patients without T2DM.

### Longitudinal impact of Type 2 Diabetes on development of clinical milestones

Over the total follow up period, MCI developed in 40 (56%) of 71 patients with PD+T2DM and 340 (34%) of 986 patients with PD. Substantial gait impairment developed in 36 (24%) of 147 patients with PD+T2DM, and in 232 (13%) in 1737 patients with PD (Figure 3). Adjusting for differences in age, sex, age, PD duration, H&Y stage, LEDD and BMI, cox proportional hazard survival analysis indicated T2DM was a predictor for patients to develop substantial gait impairment (HR 1.55, CI 1.07-2.23, p=0.020) and also MCI (HR 1.74, CI 1.19-2.55, p=0.004), compared to the PD group. There were no significant differences in the time to develop H&Y stage 3, dyskinesia, motor fluctuations, hallucinations, ICDs-RD, loss of independence, depression (Supplementary Figure 2).

**Figure 3:**
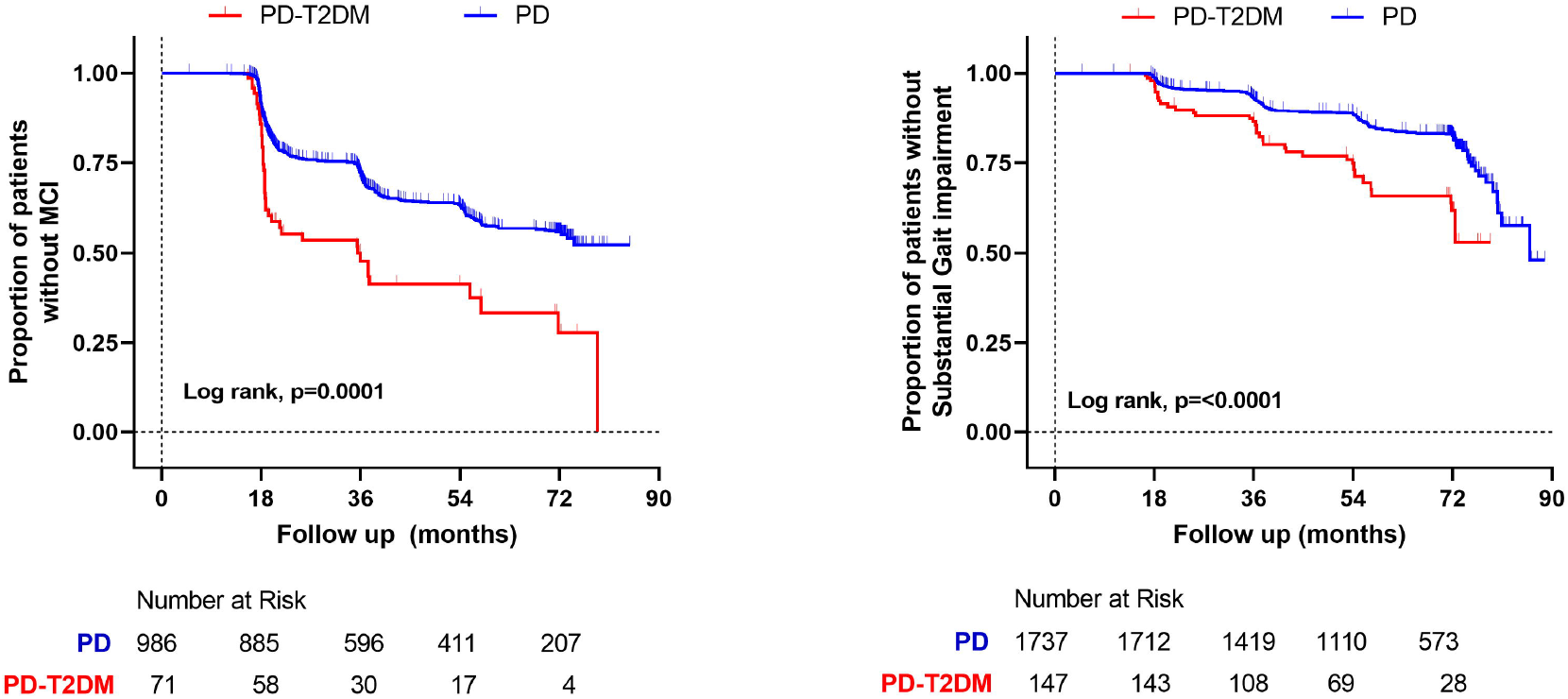
Timeline for the development of mild cognitive impairment, and substantial gait impairment, comparing PD cases with and without T2DM. Kaplan Meier curves show the significantly shorter time to develop both of these complications, in patients with T2DM.

**Figure 4:**
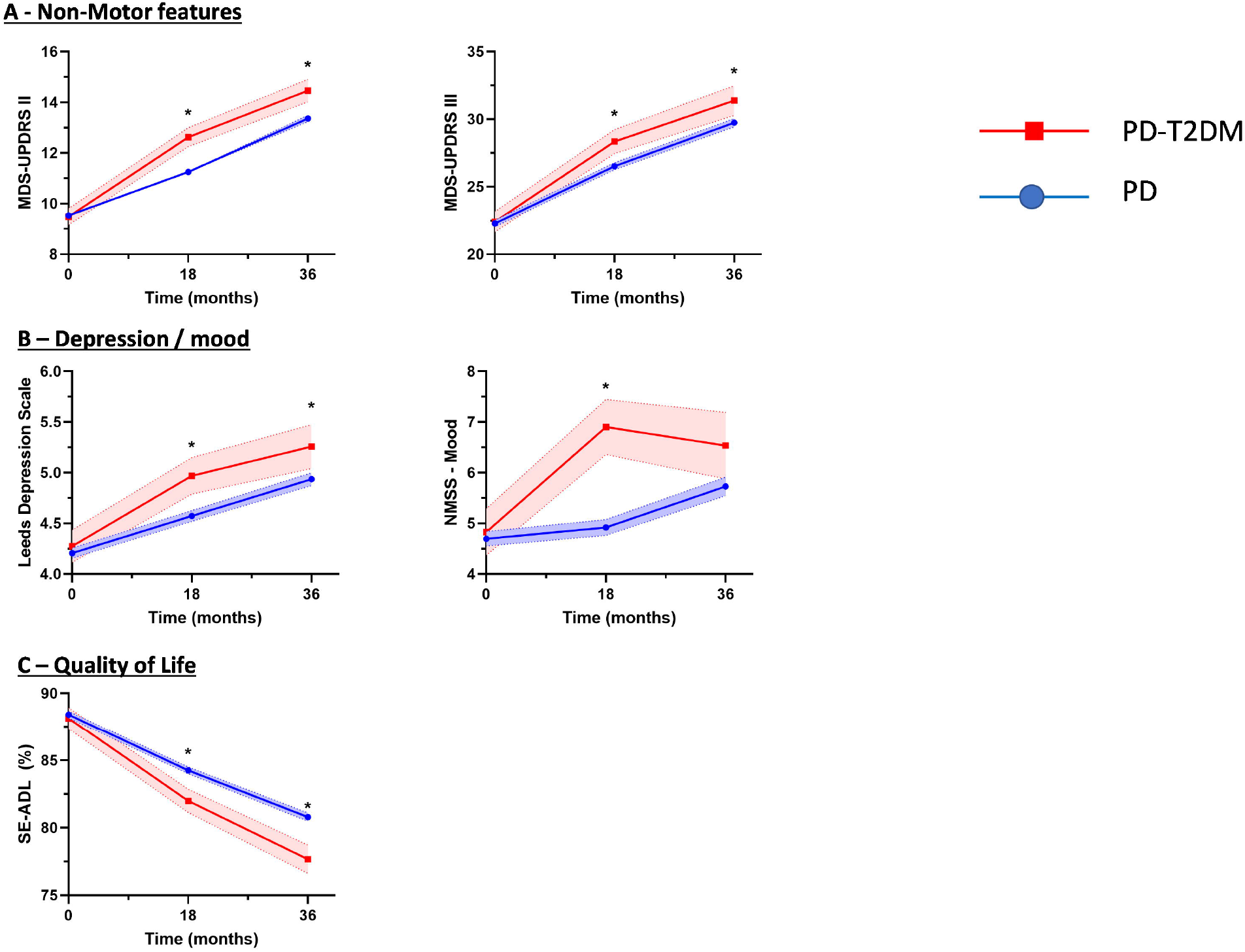
Time course of features in patients with PD, comparing those with and without T2DM. Progression was significantly faster for several domains. * denotes p<0.05; MDS-UPDRS (Movement disorders society Unified Parkinson’s disease rating scale); NMSS (Non-motor symptoms score); SE-ADL (Schwab and England Activities of Daily Living Scale)

### Longitudinal impact of Type 2 diabetes on progression of PD symptoms

Patients were followed up for a mean of 36 months. When modelling the change in motor symptoms (MDS-UPDRS II), there was a significant group × time interaction in the mixed model (P_=_0.012), after adjusting for age, sex, gender, ethnicity, baseline motor score, LED and BMI, indicating that there was a significant difference in the progression of motor symptoms in patients with T2DM. During the follow up, patients with T2DM had significantly more severe motor symptoms as reported by MDS-UPDRS II scores. In addition, this group also had significantly worse MDS-UPDRS III scores at each time point.

There was also a significant group × time interaction when modelling change in mood as measured by the NMSS Mood subscore (p=0.041), suggesting that the PD-T2DM group had a significant difference in progression of mood symptoms. Supporting this, patients in the PD+T2DM group had significantly worse depression scores at each time point (p=0.023).

Patients in the PD-T2DM group also had significantly worse quality of life scores over the follow up period compared to the PD group (p=0.001), with only the PD+T2DM group reporting loss of independence compared to the PD group (77.7% (1.0) vs 80.8% (0.3), p=0.04) after 36 months, however the group x time interaction failed to reach the conventional threshold for significance (p=0.077). There were no significant differences in motor fluctuations, or dyskinesia (Supplementary Figure 3).

### Exploratory analysis of the impact of metformin in patients with T2DM on clinical markers and progression of PD

There were no consistent effects of metformin on PD symptoms in patients recently diagnosed with PD, and metformin did not offer any benefit in slowing the development of key clinical milestones (Supplementary Table 3 and Supplementary Figures 4, 5, 6).

## Discussion

In this study we evaluated the impact of T2DM on symptom severity and disease progression in patients recently diagnosed with PD. First, at baseline entry into the study, after a disease duration of 15 months, the presence of T2DM was independently associated with more sever motor symptoms and greater overall burden of non-motor symptoms, with poorer cognitive scores as assessed by the MoCA. In addition, T2DM conferred an increased risk of patients having depression and substantial gait impairment, and despite adjusting for differences in disease severity and stage, patients with T2DM were on greater amounts of dopaminergic medication. These consistent negative effects on aspects of PD were reflected in significantly worse quality of life scores and increased levels of dependency in PD patients with T2DM compared to PD patients without T2DM. Secondly, patients with T2DM had significantly worse progression of motor symptoms over time and higher risk of developing substantial gait impairment and MCI than patients without T2DM. Finally, an exploratory analysis suggested metformin use among diabetics did not confer any protective effects on non-motor or motor symptoms or alter the time to develop clinical milestones. Overall these findings suggest T2DM contributes to more severe symptoms and alters long term outcomes in patients with PD.

Our findings that PD patients with T2DM have more aggressive disease are in keeping with earlier smaller studies that suggested even in patients recently diagnosed with PD with a disease duration of around 6-18 months, patients with T2DM have more severe motor symptoms and cognitive impairment than patients without T2DM^7,8^. The potential reason for this important association may stem from the fact that beyond sharing age as a risk factor, both T2DM and PD share several pathological processes encompassing inflammation, lysosomal dysfunction, and mitochondrial dysfunction that may lead to metabolic dysfunction or neurodegeneration. T2DM is characterised by hyperglycaemia, and it has been shown that this may increase the risk of neurodegeneration, mediated through elevated levels of advanced glycation end products (AGEs) and the subsequent impact on alpha synuclein aggregation and pro-inflammatory pathways^18^. A further hypothesis relates to the potential overlap between peripheral insulin resistance in T2DM and the development of brain insulin resistance in PD. Brain insulin signalling plays an important role in neuronal cell survival and modulates a number of cellular processes disrupted in PD including autophagy, neuroinflammation and mitochondrial function via modulation of the kinases Akt and MAPK^19^.

In addition, we found that T2DM was directly associated with significantly faster progression of motor symptoms and substantial gait impairment similar to previous reports. As impairments of gait and postural stability likely contribute to increased risk of falls and fractures in this population^20^, it is possible that intensive physiotherapy and gait re-training approaches would give the greatest benefit to these higher risk subjects^21^. One explanation for the increased risk of postural instability may relate to T2DM increasing the risk of cerebrovascular disease and cardiovascular conditions, which can lead to parkinsonism and gait disturbance in older individuals ^11^. But after adjusting for differences in cerebrovascular disease and cardiovascular conditions, T2DM remained a significant independent risk factor for the development of gait impairment, suggesting other mechanisms may also contribute. Furthermore, although other diabetic co-morbidities such as peripheral neuropathy and peripheral vascular disease may have also contributed to the accelerated gait deterioration in the PD+T2DM group, these co-morbidities are unlikely to explain the overall worsening of motor and non-motor symptom progression in the longitudinal analysis, suggesting mechanisms independent of direct diabetes related complications. Insulin receptors are widely expressed throughout the substantia nigra, and regulate dopamine synthesis and clearance. Consequently states of insulin resistance lead to reduced dopamine transporter cell surface expression and reduced synaptic dopamine signalling^22^. This is further borne out by clinical studies demonstrating PD patients with T2DM have lower striatal dopamine transporter binding and faster motor progression^8^.

Although a previous study showed that PD patients with T2DM were more likely to have cognitive impairment after a mean of 6 years disease duration, we have shown that even after a mean of 18 months, there was significantly worse cognitive test scores and a higher proportion of patients with MCI in the PD+T2DM group. In addition, we showed in patients without cognitive impairment at baseline, the PD+T2DM group were almost twice as likely to develop MCI subsequently. One possible explanation for these findings is that T2DM is a risk factor for vascular dementia, and that poorer cognitive scores may be explained by differences in the degree of leukoaraiosis. Our study did not necessitate structural imaging, so we were not able to perform a systematic analysis of this topic, but there is some support from the available imaging, as we have reported previously^23^. Conversely, others demonstrate no difference in leukoaraiosis between PD patients with and without T2DM^12^, and that microstructural changes such as white matter hyperintensities were not markedly different in patients with T2DM compared to matched aged individuals^24^. T2DM has long been established as a known risk factor for AD^25^, and furthermore, brain insulin resistance is associated with cognitive dysfunction, increased aggregation of amyloid beta, hyperphosphorylated tau, pro-inflammatory pathway activation and impaired glucose metabolism^26^. It has long been established that molecular interactions between pathological proteins may occur within the same brain in various distribution patterns, cause variable phenotypes and mixed pathologies, and so it is possible that T2DM may lead to promotion of AD pathology in a subset of PD patients, increasing the risk of developing cognitive impairment.

The present study is the first to report the impact of T2DM on non-motor symptoms in PD. At study entry, in patients with a mean disease duration of 18 months, patients with T2DM already had a greater overall non-motor symptom burden (as measured by MDS-UPDRS I and NMSS), and reported poorer sleep compared to patients without T2DM. Interestingly, the main drivers for the differences in total NMSS scores were primarily sleep, mood and memory issues – which were themselves captured on separate scales. Sleep disturbances in PD are common, comprising a spectrum of sleep disorders and often significantly contribute to poor quality of life^27^, however this study reports for the first time that T2DM may be an independent factor associated with these issues. Interestingly some studies have shown that patients with insulin resistance exhibit more sleep apnoea, insomnia, and daytime sleepiness^28^. Given the complexity of sleep disorders, it is difficult to discern underlying mechanisms for this, however significant differences remained despite adjusting for differences in BMI and depression scores (data not shown), suggesting that sleep apnoea and concurrent low mood may not account for all the changes seen. Nevertheless, the identification that patients with T2DM may be prone to poorer sleep may aid clinicians in guiding management decisions when prescribing medications that may worsen sleep (such as dopamine agonists). We also report the first association of T2DM with mood in PD. T2DM was associated with a significantly worse mood scores in patients assessed at entry into the study, and patients reported consistently lower mood throughout follow up than the PD group. Furthermore, T2DM was independently and directly associated with worse longitudinal progression of the NMSS mood subscores throughout the follow up period. Accumulating evidence suggests insulin resistance is a risk factor for depression^29–31^, with increased prevalence of depression in patients with T2DM^32^, and this is supported by our findings. Although risk factors for depression in PD have been explored, T2DM has not previously been identified as a risk factor for this important comorbidity^33^, and this association may aid clinicians in identifying patients at increased risk of depressive disturbances. This is also the first report that suggests a link between T2DM and dopamine dysregulation. Contrary to other non-motor symptoms, patients with T2DM reported fewer symptoms of ICDs-RD (classified as related behaviours that have a contrasting clinical presentation with respect to the four major ICDs and include punding, hobbyism and aimless wandering^34^).The pathophysiology of ICDs-RD like punding is complex, but is thought to involve stimulation of D1 and D2 receptors, and studies in animals support the hypothesis that the reward system acts by means of increasing dopamine in the nucleus accumbens and the dorsal striatum (becoming conditioned cues)^35^. Interestingly, insulin signalling has a reciprocal relationship to dopamine action and impacts behaviours such as reward and mood, and clinical studies have shown insulin resistance is associated with less endogenous dopamine at D2/3 receptors^36^, thus patients with T2DM may be at lower risk of these behaviours. Given the interest in anti-glycaemic drugs as potential novel treatments for PD, an exploratory analysis was performed to evaluate if metformin could reduce or restore the negative impact of T2DM. The present study demonstrated that metformin use did not confer any protective effects on the diabetic population on any motor, non-motor or quality of life outcomes, and in fact MoCA scores were significantly lower (worse) at 36 months in the PD+T2DM/Met group. Metformin is one of the most commonly used diabetic drugs and acts an insulin sensitizer via activation of the AMPK pathway. Interest in metformin as a potential neuroprotective drug is supported by in vivo and in vitro studies demonstrating metformin can restore dopaminergic dysfunction and reduce aggregation of alpha-synuclein^37^. However, results from other studies are conflicting. Studies have shown metformin increases the risk of developing dementia and PD^38,39^, and can exacerbate intra- and extra-cellular production of amyloid-beta^40^. Collectively, our data may imply that metformin does not offer additional benefit on neurological outcomes in diabetes but a more robust study is needed to better inform on this issue. Of note, due to insufficient numbers of patients prescribed the newer classes of glucagon-like peptide-1 (GLP-1) drugs, we were unable to perform this analysis

The main strength of our study is that this is a large and longitudinal study of PD and patients were well phenotyped with a variety of PD scales to characterise the severity of a number of non-motor and quality of life scales, in addition to the typical motor scales, allowing us to gain a global overview of PD severity, as well as individual symptoms. The prevalence of T2DM in our PD cohort was approximately 10% - which is in line with other reported studies, and thus the large number of patients with T2DM allowed tentative casual inferences to be made and some generalizability of the findings. A large amount of demographic and other data was readily available, and so by utilising these data and including our DAG allowed us to select a set of covariates (based on a literature review and expert opinion) that allowed the estimation of causal effects from observed data and were helpful in delineating and understanding confounders and potential sources of bias, and examining the independent effects of T2DM in sequential analyses.

An important limitation in this study, as is observed in many other longitudinal studies of this nature, is the drop-out of patients during follow up. As this may introduce a bias regarding surviving patients, a decision was made to only include follow up data for the first 36 months for one aspect of the longitudinal analysis. The survival analyses curves may have been influenced by censored data and further studies will be needed to confirm these findings. T2DM was identified using medication data and self-report, and clinical / serological data was unavailable, suggesting that diabetes may have been under recognized in this study. However, this typically would have weakened our ability to detect an association between PD and T2DM. In addition, this study was not able to fully examine different facets of T2DM phenotypes, such as the effect of severity of diabetes, duration of disease, and other comorbidities that may have impacted on the relation of T2DM to PD severity. Particularly challenging is controlling for changes to other non-PD and diabetic medication made throughout the follow up. In conclusion, we have identified T2DM is an independent risk factor associated with more severe motor symptoms, non-motor symptoms, and poorer quality of life scores in patients recently diagnosed with PD. Furthermore, T2DM is identified as a new factor that contributes to faster motor and non-motor symptom progression, and increases the risk of developing MCI and gait impairment. The importance of this is that insulin resistance is potentially a modifiable metabolic state, with multiple peripheral and central targets for intervention, and thus represents a novel target for ameliorating parkinsonian symptoms and neurodegeneration, and progression to disability and dementia. Targeting insulin resistance to treat PD is supported by recent cohort studies showing that better managed diabetic control is associated with less severe PD symptoms and disease progression^7^, while diabetic patients on DPP-IV inhibitors have less nigrostriatal dopamine degeneration and better long-term motor outcomes in diabetic patients with than those not treated with this class of drug^41^. Multiple trials of anti-glycaemic medications for the treatment of PD and other neurodegenerative diseases are currently underway and the results of these will greatly inform the next generation of novel PD treatments.

## Data Availability

All data produced in the present work are contained in the manuscript

