## Supplemental Material for "The impact of Type 2 diabetes in Parkinson’s disease"

**Supplementary Material**

**Supplementary Methods:**

Outcomes included appearance of motor fluctuations, dyskinesias, impulse control disorders, dopamine dysregulation, and hallucinations. These outcomes were regarded as binary endpoints, corresponding to MDS-UPDRS item responses of ≥1, as per previous studies^1^. In addition, the onset of cognitive decline was defined as the development of mild cognitive impairment (MCI). We used MDS level I criteria^2^ to define MCI, which has been shown to have prognostic value in identifying patients at risk of developing PDD^3^, and used Montreal Cognitive Assessment (MoCA) test scores to categorise cases into normal (MoCA≥26); and MCI (MoCA 21-25, with no functional cognitive impairment as assessed by MDS-UPDRS 1.1); matching previous studies^4,5^. Substantial gait impairment was defined as score >3 on MDS-UPDRS III Q10 (requiring the use of assistance to mobilise). The Non-Motor Symptoms Scale (NMSS) was used to derive non-motor symptom burden. Loss of independence is an important determinant of quality of life^6^, and has been defined previously as Hoehn & Yahr stage >3^7^, and Schwab and England ADL scale <80% ^8^ and we used both of these scores to define loss of dependence. The Questionnaire for impulsive-compulsive disorders in PD (QUIP) was used to define impulse control disorders (ICD) and ICDs-RD Individuals with ICD were defined as any affirmative response to questions pertaining to pathological gambling, hypersexuality, binge eating and compulsive buying; and ICDs-RD were defined as individuals with any affirmative response to questions regarding punding, hoarding, walkabout/aimless wandering, which has previously been shown to have good sensitivity and specificity for identifying these disorders^9^. Depression was defined using the Leeds Anxiety and Depression Scale (LADS) with a cut-off score >6^10^. To conduct our exploratory analysis, medication lists were used to identify patients treated with anti-diabetic medication. Metformin was the most commonly prescribed drug, which directed our exploratory analysis (too few patients were prescribed the newer glucagon-like peptide-1 (GLP-1) class of drugs to perform an analysis).

**Supplementary Figure 1:**

Directed acyclic graph illustrating confounding and mediating factors to determine the causal impact of T2DM on PD severity. This considers each variable in relation to the exposure and outcome, as both the failure to adjust for a confounder, and over-adjusting for an intermediate variable can lead to biased results^11,12^. Included associations were based on past literature and expert knowledge, and we used the program DAGitty^13^, which uses an algorithm to identify a “minimally sufficient adjustment set” containing no redundant variables, to allow us to adjust models for confounders and make causal inferences.

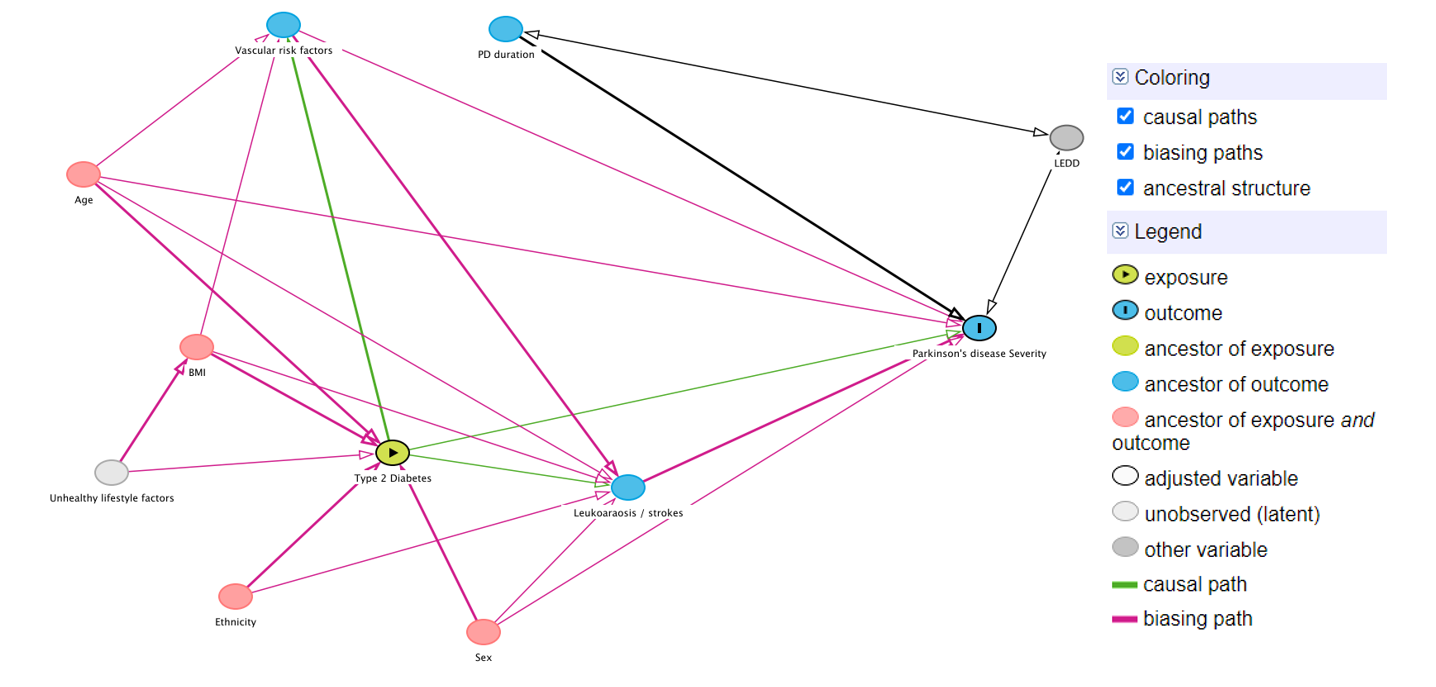

**Supplementary Table 2:**

Longitudinal follow up. Univariate and multivariate Cox regression analyses for the development of each disease marker / clinical outcome in PD according to the presence of T2DM. Multivariate analyses adjusted for age, sex, ethnicity, disease duration, Hoehn & Yahr stage, BMI and LEDD.

| Disease marker / clinical milestone | Univariate  HR (95% CI) | p value | Multivariate  HR (95% CI) | p value |
| --- | --- | --- | --- | --- |
| **Substantial gait impairment**  *(MDS-UPDRS 3·10>3)* | 2·33 (1·64-3·32) | <0·0001 | 1·55 (1·07-2·23) | 0·020 |
| **MCI**  *(MoCA<26)* | 2·16 (1·56-3·01) | <0·0001 | 1·74 (1·19-2·55) | 0·004 |
| **Hallucinations**  *(MDS-UPDRS 1·2>1)* | 0·91 (0·68-1·21) | 0·503 | 0·80 (0·59-1·08) | 0·139 |
| **Depression**  *(LADS>6)* | 1·42 (1·0-2·02) | 0·051 | 1·32 (0·92-1·90) | 0·134 |
| **ICD**  (QUIP 1-4 >1) | 1·13 (0·79-1·61) | 0·494 | 1·13 (0·78-1·65) | 0·524 |
| **Dyskinesia**  *(MDS-UPDRS 4·1>1)* | 1·12 (0·73-1·71) | 0·608 | 1·08 (0·67-1·73) | 0·748 |
| **Motor fluctuations**  *(MDS-UPDRS 4·3>1)* | 1·23 (0·91-1·65) | 0·185 | 1·25 (0·91-1·72) | 0·167 |
| **Loss of independence**  *(SE-ADL<80%)* | 1·56 (1·15-2·10) | 0·004 | 1·18 (0·89-1·61) | 0·289 |
| **Loss of independence**  (H&Y >3) | 1·56 (1·13-2·15) | 0·007 | 1·11 (0·79-1·56) | 0·548 |

MCI (Mild cognitive impairment); ICD (impulse control disorder); MDS-UPDRS (Movement Disorders Society Parkinson’s Disease Rating scale); H&Y (Hoehn & Yahr stage)

**Supplementary Figure 2:**

Kaplan Meier curves - no significant differences were observed in time to develop H&Y Stage 3, Dyskinesia, Motor fluctuations, Hallucinations, ICD, Loss of independence, Depression

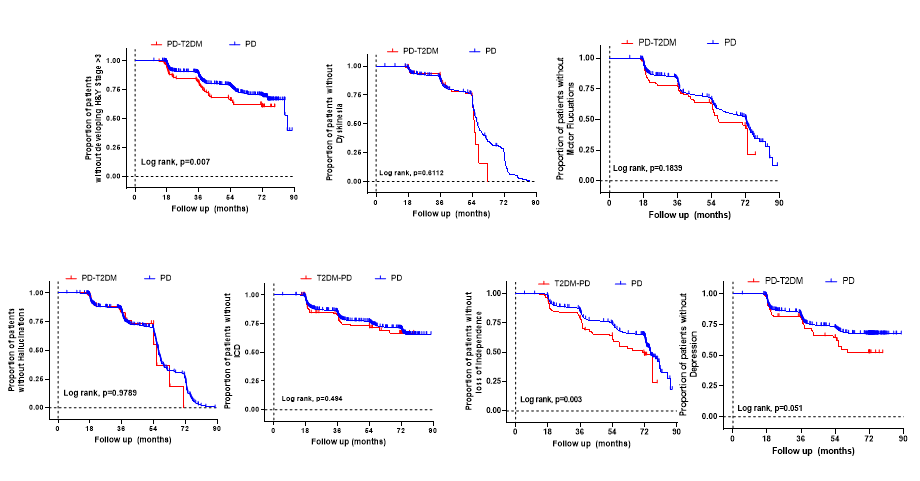

MCI (Mild cognitive impairment); ICD (impulse control disorder); MDS-UPDRS (Movement Disorders Society Parkinson’s Disease Rating scale); H&Y (Hoehn & Yahr stage)

**Supplementary Figure 3:**

Linear mixed modelling progression of symptoms of PD

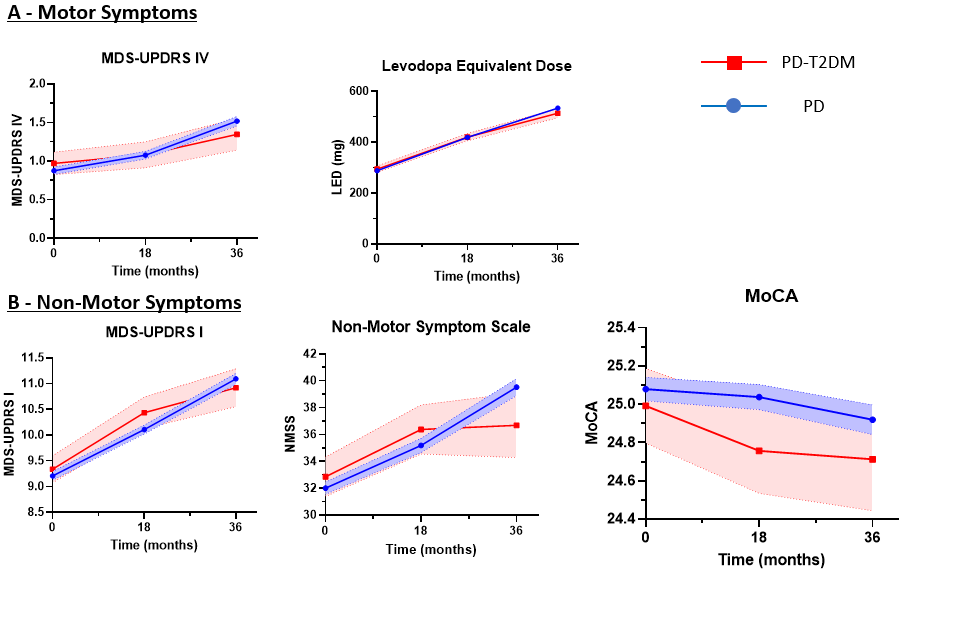

**Supplementary Table 3:**

Clinical Features of patients within 18 months of diagnosis of PD compared to patients with co-morbid T2DM and patients treated with metformin

| **Demographics** |  | | | **PD**  **(n=1736)**  **Mean (SE)*** | **PD-T2DM**  **(n=72)**  **Mean (SE)*** | **PD-T2DM/Met**  **(n=95)**  **Mean (SE)*** | **P value** |
| --- | --- | --- | --- | --- | --- | --- | --- |
|  | Age | | | 67·19 (0·2) | 72·26 (0·9) | 70·11 (0·9) |  |
|  | Age of diagnosis | | | 65·85 (0·2) | 70·85 (0·9) | 68·84 (0·9) |  |
|  | Disease duration, years | | | 1·33 (0·1) | 1·41 (0·1) | 1·27 (0·1) |  |
|  | Sex, males (%) | | | 1137 (90·4) | 51 (69·9) | 70 (74·5) | 0·098 |
|  | Ethnicity, white (%) | | | 35 (2·0) | 1 (1·4) | 1 (1·1) | 0·761 |
|  | BMI | | | 26·74 (0·1) | 29·68 (0·6) | 29·67 (0·63) |  |
| **Aspect of PD** | **Scale** | | |  |  |  |  |
| **Non-motor symptoms** | **UPDRS I** | | | **9·18 (0·1)** | **11·1 (0·6)** | **9·98 (0·5)** | **0·006^a,b^** |
|  | **NMSS Total** | | | **31·69 (0·7)** | **39·4 (3·5)** | **37·28 (3·1)** | **0·030** |
|  | **Leeds Anxiety Index** | | | **4·10 (0·8)** | **4·52 (0·4)** | **4·7 (0·3)** | **0·149** |
|  |  | *Anxiety (LAD>6), n (%)* | | *392 (23·1)* | *18 (26·5)* | *29 (33·3)* | *0·078* |
|  | **Leeds Depression Index** | | | **4·22 (0·7)** | **5·06 (0·3)** | **5·03 (0·3)** | **0·009** |
|  |  | *Depression (LAD>6, n (%)* | | *380 (22·4)* | *22 (31·4)* | *33 (37·1)* | *0·002* |
| **Sleep** | **PDSS** | | |  |  |  |  |
|  | **ESS** | | | 6·71 (0·10) | 8·15 (0·5) | 7·56 (0·5) | 0·010^a^ |
| **Cognition** | **MOCA Total** | | | 25·08 (0·1) | 23·97 (0·4) | 23·33 (0·3) | <0·001^a,c^ |
|  |  | *MCI (MOCA<26), n (%)* | | 788 (48·2) | 39 (60·0) | *57 (67·1)* | 0·001 |
| **Psychiatric co-morbidity** |  | *Dopamine dysregulation* | | *37 (2·1)* | *4 (5·6)* | *2 (2·1)* | *0·156* |
|  |  | *ICD (QUIP>1·0)* | | *43 (2·6)* | *3 (4·3)* | *4 (4·7)* | *0·360* |
|  |  | *Hallucinations* | | *43 (2·5)* | *5 (6·9)* | *3 (3·2)* | *0·065* |
| **Motor symptoms** | **UPDRS II** | | | 9·71 (0·1) | 11·01 (0·7) | 9·84 (0·6) | 0·246 |
|  | **UPDRS III** | | | 22·50 (0·3) | 26·6 (1·5) | 25·1 (1·2) | 0·006^a^ |
|  |  | *Substantial gait impairment, n (%)* | | *46 (2·6)* | *9 (12·7)* | *11 (16·7)* | *<0·001* |
|  | **UPDRS IV** | | | 0·73 (0·1) | 0·8 (0·2) | 0·9 (0·2) | 0·625 |
|  |  | *Dyskinesia, n (%)* | | 69 (4·0) | 2 (2·8) | *5 (5·4)* | 0·696 |
| **Quality of life** | **PDQ8 Total** | | | 5·77 (0·1) | 6·78 (0·5) | 6·0 (0·4) | 0·213 |
|  | **EQ5D VAS** | | | 77·2 (0·4) | 70·1 (2·0) | 73·0 (1·7) | <0·001^a^ |
|  | **EQ5D Index** | | | 0·72 (0·1) | 0·66 (0·2) | 0·68 (0·1) | 0·004^a^ |
|  | **SE-ADL** | | | 88·5 (0·2) | 83·4 (1·3) | 86·4 (1·1) | <0·001^a^ |
|  |  | *Loss of independence, n (%)* | | 150 (8·6) | 19 (26·8) | 14 (15·1) | <0·001 |
|  |  | *Hoehn & Yahr >3, n (%)* | | 107 (6·2) | 13 (18·1) | 10 (10·8) | <0·001 |
| **Medication** | **Levodopa Equivalent dose, mg** | | | 290·20 (4·6) | 281·23 (23·1) | 348·34 (20·4) | 0·016^b^ |
|  |  | *Untreated, n (%)* | |  |  |  |  |

^a^ Difference between PD-T2DM and PD

^b^ Difference between PD-T2DM and PD-T2DM/Met

^c^ Difference between PD and PD-T2DM/Met

MCI (Mild cognitive impairment); ICD (impulse control disorder); MDS-UPDRS (Movement Disorders Society Parkinson’s Disease Rating scale); H&Y (Hoehn & Yahr stage)

**Supplementary Figure 4:**

Longitudinal impact of T2DM on symptoms in PD per group.

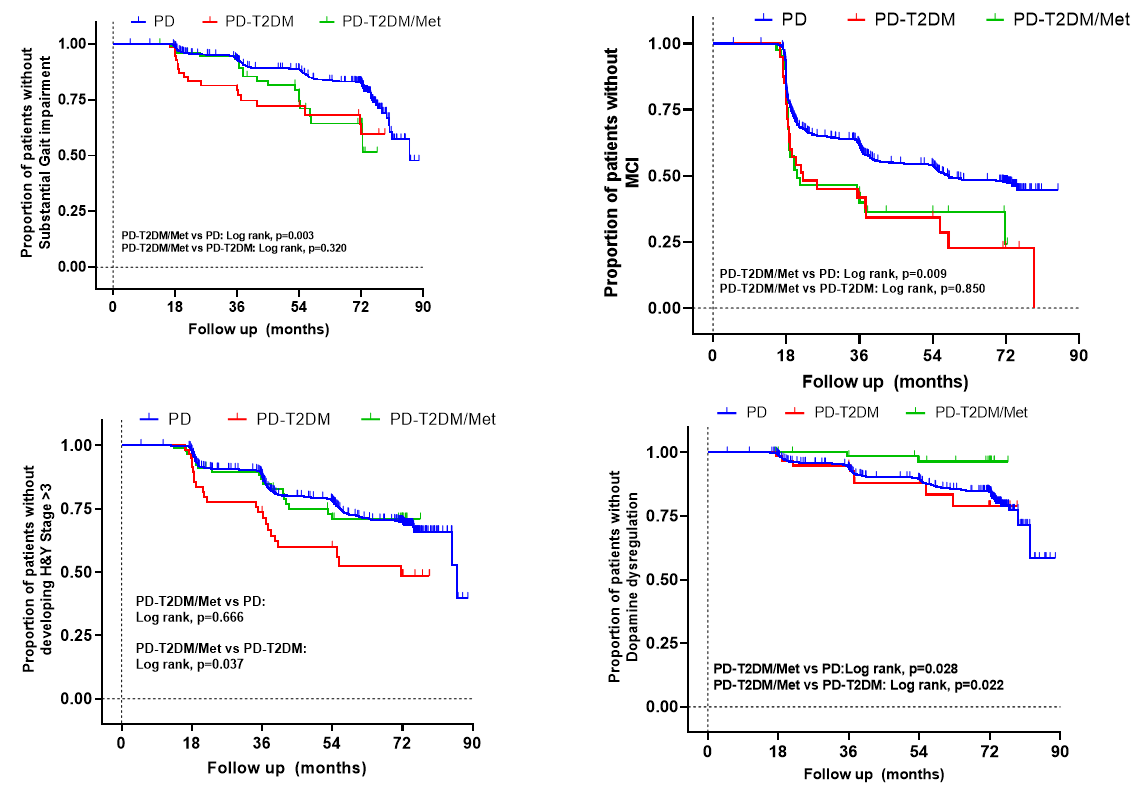

**Supplementary Figure 5:**

Longitudinal impact of T2DM on symptoms in PD per group.

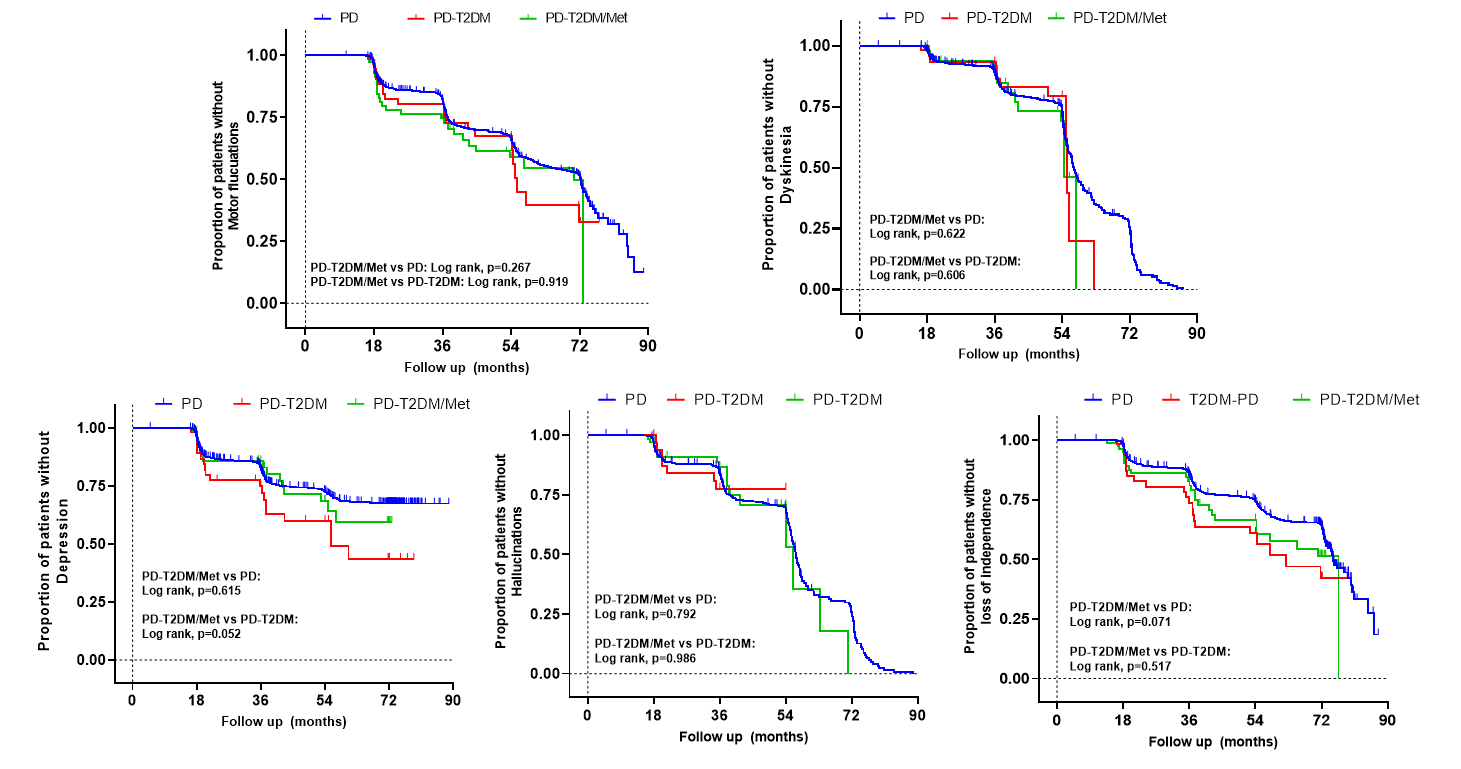

**Supplementary Table 3:**

Multivariate Cox regression analyses for the development of each disease marker / clinical outcome in patients with PD according to the use of metformin. Multivariate analyses adjusted for age, sex, ethnicity, disease duration, Hoehn & Yahr stage, BMI and LED

| Disease marker / clinical milestone | PD-T2DM/Met  vs  PD-T2DM | | PD-T2DM/Met  Vs  PD | |
| --- | --- | --- | --- | --- |
|  | Multivariate  HR (95% CI) | p value | Multivariate  HR (95% CI) | p value |
| **Substantial gait impairment**  *(MDS-UPDRS 3·10>3)* | 1·06 (0·48-2·34) | 0·890 | 1·212 (0·94-1·56) | 0·137 |
| **MCI**  *(MoCA<26)* | 0·91 (0·49-1·68) | 0·768 | 1·18 (0·96-1·45) | 0·112 |
| **Development of H&Y >3** | 0·62 (0·31-1·20) | 0·157 | 0·89 (0·70-1·13) | 0·370 |
| **Hallucinations**  *(MDS-UPDRS 1·2>1)* | 0·70 (0·23-2·15) | 0·538 | 0·98 (0·74-1·29) | 0·908 |
| **Depression**  *(LADS>6)* | 1·34 (0·64-2·82) | 0·426 | 1·34 (1·06-1·71) | 0·015 |
| **ICD**  (QUIP 1-4 >1) | 0·65 (0·30-1·39) | 0·266 | 0·97 (0·75-1·25) | 0·833 |
| **Dyskinesia**  *(MDS-UPDRS 4·1>1)* | 1·32 (0·38-4·53) | 0·654 | 1·05 (0·76-1·43) | 0·740 |
| **Motor fluctuations** | 1·17 (0·59-2·30) | 0·654 | 1·12 (0·91-1·38) | 0·260 |
| **Loss of independence**  *(SE-ADL<80%)* | 1·18 (0·93-3·53) | 0·080 | 1·08 (0·89-1·32) | 0·406 |

**Supplementary Figure 6:**

Linear mixed modelling progression of symptoms of PD vs PD+T2DM and PD+T2DM/Met

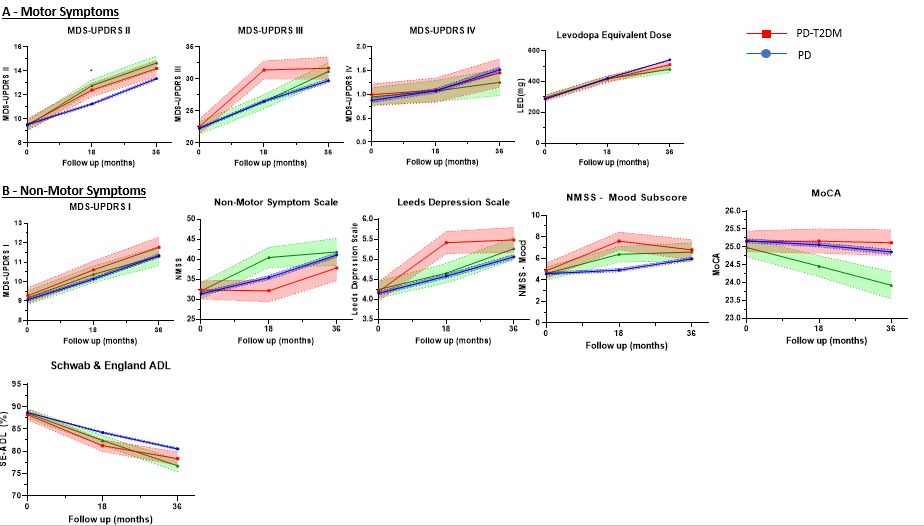
